# Is the glass half full? Extraction-free RT-LAMP to detect SARS-CoV-2 is less sensitive but highly specific compared to standard RT-PCR in 101 samples

**DOI:** 10.1101/2020.12.07.20239558

**Authors:** John J. Schellenberg, Margaret Ormond, Yoav Keynan

**Author notes:** Corresponding author: J. Schellenberg. **Statement Regarding Conflict of Interest:** The authors declare no conflicts of interest. **Author contributions:** JJS: Conceptualization, Methodology, Investigation, Writing - Original draft preparation. MO: Conceptualization, Writing - Reviewing and Editing. YK: Conceptualization, Supervision, Writing – Reviewing and Editing.

## Abstract

The current scale of public and private testing cannot be expected to meet the emerging need for higher levels of community-level and repeated screening of asymptomatic Canadians for SARS-CoV-2. Rapid point-of-care techniques are increasingly being deployed to fill the gap in screening levels required to identify undiagnosed individuals with high viral loads. However, rapid, point-of-care tests often have lower sensitivity in practice. Reverse transcription loop-mediated isothermal amplification (RT-LAMP) for SARS-CoV-2 has proven sensitive and specific and provides visual results in minutes. Using a commercially available kit for RT-LAMP and primer set targetting nucleocapsid (N) gene, we tested a blinded set of 101 archived nasopharyngeal (NP) swab samples with known RT-PCR results. RT-LAMP reactions were incubated at 65°C for 30 minutes, using heat-inactivated nasopharyngeal swab sample in viral transport medium, diluted tenfold in water, as input. RT-LAMP agreed with all RT-PCR defined negatives (N=51), and all positives with Ct less than 20 (N=24), 65% of positives with Ct between 20-30 (N=17), and no positives with Ct greater than 30 (N=9). RT-LAMP requires fewer and different core components, so may not compete directly with the mainline testing workflow, preserving precious central laboratory resources and gold standard tests for those with the greatest need. Careful messaging must be provided when using less-sensitive tests, so that people are not falsely reassured by negative results – “glass half empty” – in exchange for reliable detection of those with high levels of virus within an hour, using <$10 worth of chemicals – “glass half full”.

## Introduction

In an unprecedented scientific feat, nucleic acid amplification tests (NAAT) for SARS-CoV-2 were published 21 days after the Chinese communicable disease control team arrived in Wuhan on 31 December 2019 ^**1**^, based on complete genome sequences published 10 days earlier ^**2**^. Hundreds of molecular diagnostic tests for SARS-CoV-2 have been introduced since then ^**3-5**^. The main diagnostic targets rely on specific host antibodies, viral proteins and viral RNA, each with its own specific benefits and limitations in accurately detecting the SARS-CoV-2 infectious period to prevent its spread. The best, most sensitive examples of tests to detect each target require high levels of laboratory expertise and specialized facilities that massively increase expense and turnaround time, especially when challenged with recent, unprecedented testing volumes. In addition, high throughput platforms used in laboratories have been diverted to testing for SARS-CoV-2 at the expense of other infectious disease testing ^**6**^.

Advice on whether to test people “without symptoms” (which represents a range of pre-symptomatic, peri-symptomatic, sub-symptomatic and truly asymptomatic phenotypes) wavers as much over time as actual demand for testing by people “without symptoms” ^**7,8**^. Constant demand on the testing sites has led to “covid fatigue”, exhaustion of resources and delays in testing for sexually transmitted infections that have not diminished during the current pandemic. The arguments for not testing individuals “without symptoms”, since only a small percentage test positive, make sense when investing the most precise, most expensive and most time-consuming test available, for such a small return, considering the cost for human and reagent resources. In a parallel system for screening people “without symptoms” (for example, the Iceland model ^**9**^), that is much cheaper and faster per person, doesn’t occupy the commercial platforms used in diagnostic laboratories, and with a 50-75% sensitivity might start looking like “a glass half full”.

Point-of-care assays that can rapidly indicate infection outside the laboratory have been approved for use in many jurisdictions. These include rapid antibody-detecting cassettes that use serum or a few drops of peripheral blood, rapid antigen tests on nasopharyngeal samples, and all-in-one qRT-PCR platforms such as Cepheid’s GeneXpert, which provides highly sensitive and specific results relative to the gold standard in a fraction of the time ^**10,11**^. However, a key limitation is lack of scalability, with each (very expensive) machine only able to run one or a few samples at a time, with only a fixed number of single-source, single-use cassettes available per site per day. The intense demand for cartridges across all jurisdictions makes overall access limited.

More recently, nucleic acid isothermal amplification (NAIA) techniques, including loop-mediated isothermal amplification (LAMP), recombinase-polymerase amplification (RPA) and transcription-mediated amplification (TMA) have been shown to detect SARS-CoV-2 with very high sensitivity and specificity ^**12,13**^. Isothermal amplification underlies several tests recently authorized for emergency use, including the ID NOW from Abbott (recently approved in Canada) and CRISPR-based detection using SHERLOCK ^**14**^ (currently not approved for sale outside the United States).

The ID NOW compares favourably to GeneXpert ^**15**^, but also shares its disadvantages in terms of single sample per use and single-source cartridges. The LAMP/CRISPR test requires no special machines or cartridges, requiring only strip tubes or plates and p10 filter tips, hence it can easily be scaled up to conduct hundreds or thousands of tests. New England Biolabs’ Colorimetric WarmStart LAMP kit can also be scaled and, thankfully, is available in Canada in various formulations. Incubation requires a simple heating block, or even a warm thermos that can hold the required temperature until DNA amplification is complete. Since DNA amplification produces protons, the pH of the reaction drops and can conveniently be interpreted by eye using colour-sensitive pH stains, such as cresol red. In this study, we evaluated how a commercial colorimetric RT-LAMP kit combined with published SARS-CoV-2 primer sets and heat-treated, diluted sample compares with “gold standard” RT-PCR analysis conducted by our central laboratory.

## Materials and Methods

### Archived sample set

A total of 101 leftover nasopharyngeal swab samples in viral transport medium, previously tested for SARS-CoV-2 by RT-PCR using E gene primers and purified RNA template on the Cobas or Panther Fusion platforms, were used to validate a recently described RT-LAMP assay. 50 samples were RT-PCR positive, with fluorescence threshold values between 10 and 37 cycles, and 51 samples were RT-PCR negative. Samples were de-identified and only RT-PCR and RT-LAMP results were retained for analysis. All study procedures were reviewed and approved by the Biomedical Research Ethics Board, University of Manitoba.

### Sample processing and heat treatment

After assigning blinded sample identifiers to reduce potential bias in interpretation, samples were thawed and briefly spun down in a mini-centrifuge to collect cells and debris. An aliquot of 60 μl, drawn from the bottom of the tube near the pelleted material, was transferred to a 1.7 ml Eppendorf tube, labelled with the blinded code, then incubated in a heating block at 95°C for 5 minutes. Serial tenfold dilutions of sample were prepared in nuclease-free water (New England Biolabs, Whitby, ON).

### RT-LAMP assay and controls

RT-LAMP was carried out using the Colorimetric WarmStart LAMP Kit (New England Biolabs) and a published LAMP primer set targeting the N gene of SARS-CoV-2, previously shown to have high sensitivity ^**16,17**^ (Integrated DNA Technologies, Coralville, IA). A 10X mixture of the six primers in the set was prepared by first suspending all primers separately in nuclease-free water at a concentration of 100 μM, then combining 16 μl each of FIP/BIP, 4 μl each of LF/LB, 2 μl each of F3/B3, and 56 μl nuclease-free water (final volume 100 μl). Serial tenfold dilutions in nuclease-free water of SARS-CoV-2 or MERS-CoV DNA (Integrated DNA Technologies) were used as positive and negative controls, respectively, and included in every test batch. Reactions were set up in a final volume of 20 μl (10 μl 2X Master Mix, 2 μl 10X primer mix, 6-7.5 μl nuclease-free water and 0.5-2 μl sample, either diluted or undiluted) and incubated in a heat block at 65°C for 30 minutes. After sitting on ice for a few minutes to sharpen contrast, colour was assessed visually and photographed. Bright yellow colour indicates a positive result, while magenta indicates a negative result. Tests with orange or pink colour were considered ambiguous and re-tested.

### Data analysis

After categorization as either positive (yellow) or negative (magenta), samples were unblinded and compared. Sensitivity, specificity and positive/negative predictive values with confidence intervals were calculated using standard formulas with the help of MedCalc: https://www.medcalc.org/calc/diagnostic_test.php.

## Results

### Sensitivity, specificity and predictive values

Sensitivity of the RT-LAMP assay using raw, heat-inactivated sample was 77% compared to RT-PCR of purified RNA extracts (Table 1). Samples with the lowest Ct values (<22) were all bright yellow by RT-LAMP, even when diluted up to 1000-fold, indicating a similar range of detectable concentration to the positive control. All RT-PCR negatives were also RT-LAMP negative (Table 1). This indicates that RT-LAMP may be most useful to more quickly detect those with high viral loads in their specimen who are most likely to transmit the virus to others.

**Table 1:**
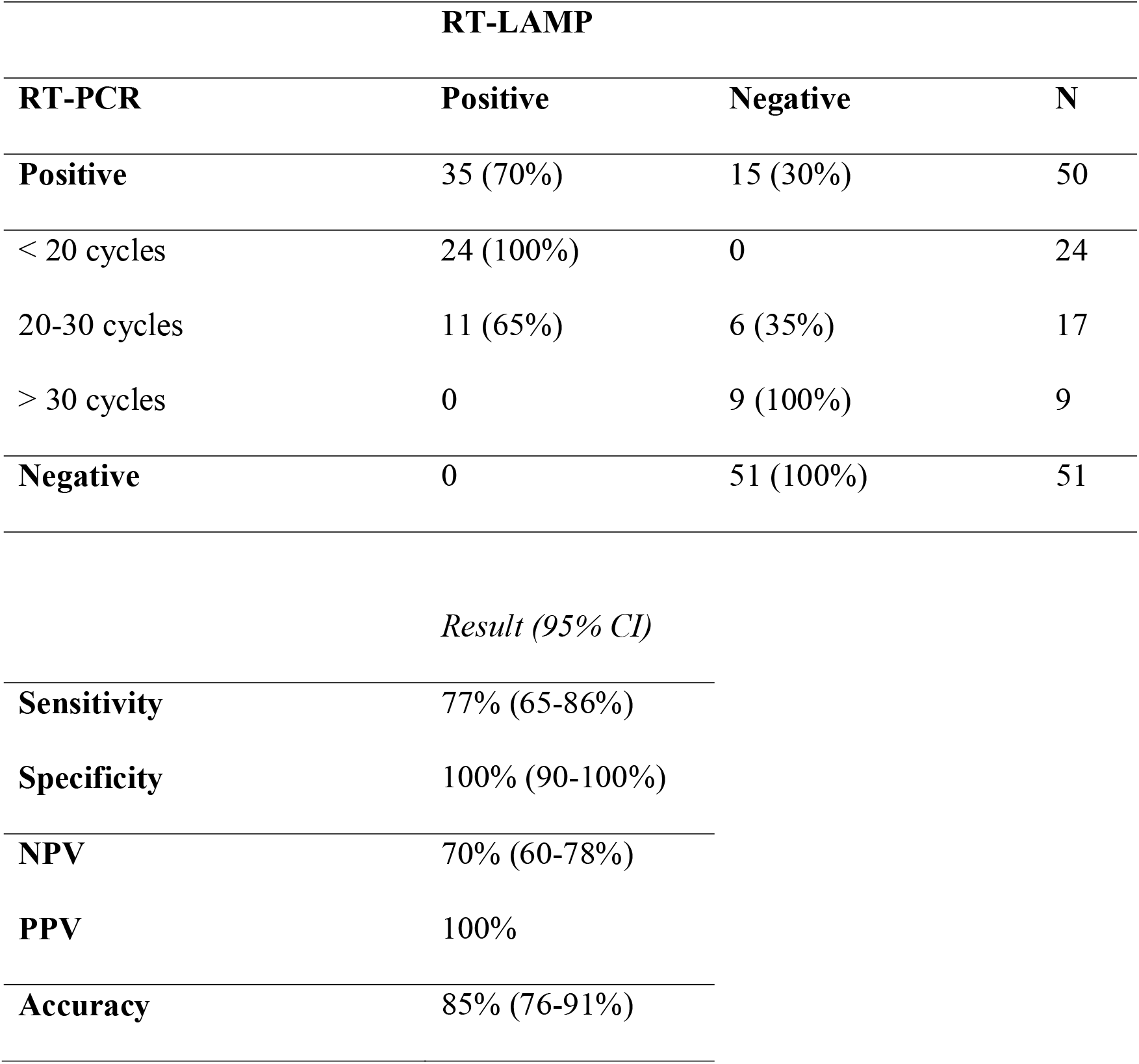
Sensitivity, specificity and predictive values of RT-LAMP with raw sample input, compared to standard RT-PCR diagnosis.

### Interpretation of colour change

Only samples that were bright yellow after 30 minutes incubation were considered positive (Figure 1). In some cases, a partial colour change resulted, from magenta to pink to orange, but did not become yellow within the period of observation in which the positive control always turns yellow (20-30 minutes, regardless of dilution factor down to 1,000 copies). This partial colour change may indicate a weak positive (all samples do eventually turn yellow), a failed reaction (not indicating anything about the sample itself), or a true negative, depending on the reason for the weak change in pH. Pink and orange reactions were also more frequent when undiluted raw sample or 10% v/v template/reaction mixture was used.

**Figure 1:**
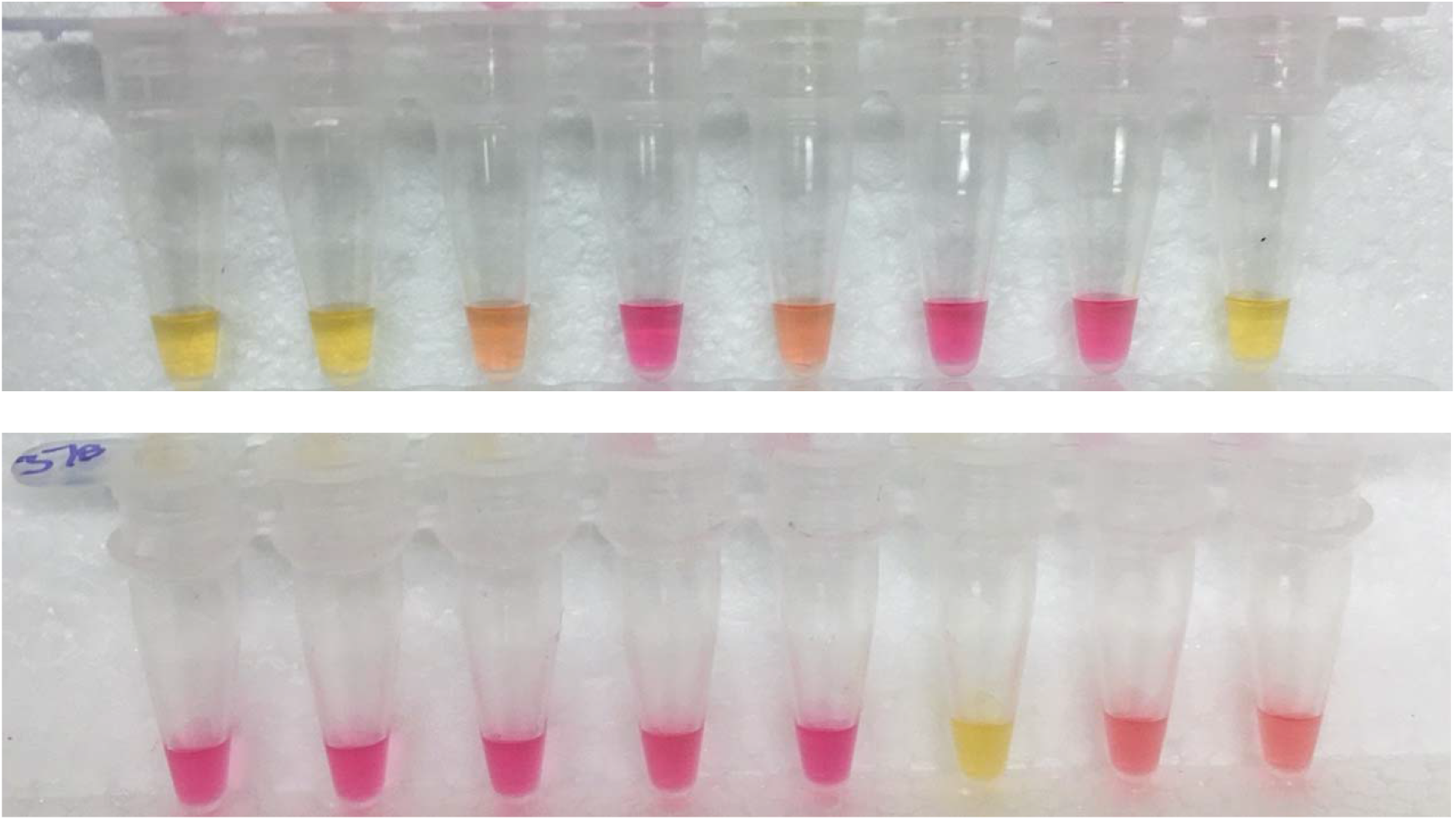
Interpretation of colour in tests on Samples 9-24. Only bright yellow results after 30 minutes at 65°C are considered positive. Magenta colour indicates a “true” negative (insufficient viral RNA), while orangey colour is considered “indeterminate” and re-tested.

### Controls and limit of detection

Negative controls were reliably magenta throughout the experiments (Figure 2) but did eventually get contaminated. Routine cleanup with DNAse and careful separation of pre-amplification and amplification/post-amplification areas are required (unless you use NEB’s more expensive dUTP kit) to prevent the massive DNA contamination that LAMP is prone to. The MERS DNA negative control at 1,000 and 10,000 copies did not produce any yellow colour, however 100,000 copies resulted in a positive reaction (not shown). SARS-CoV-2-N DNA positive control produced a bright yellow reaction within 30 minutes down to 1,000 copies, but remained magenta at lower dilutions, indicating 10^3^ viral copies per μl as the lower limit of detection for the current conditions (Figure 2).

**Figure 2:**
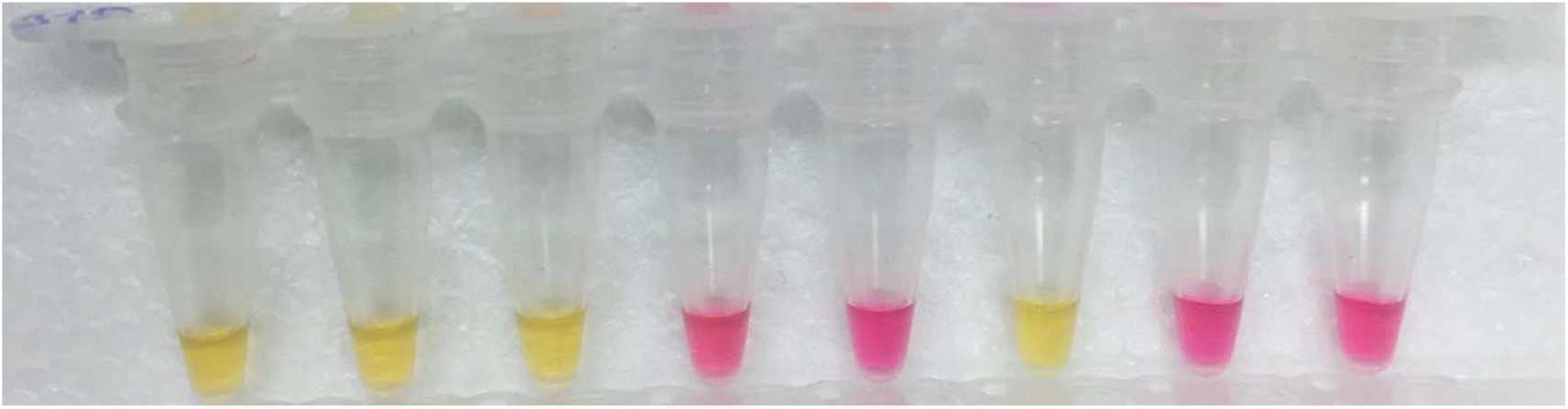
Positive and negative controls. Tubes 1-5: Tenfold dilutions of a new batch of SARS-CoV-2-N DNA (positive control), 10^5^/10^4^/10^3^/10^2^/10^1^ per μl. Tube 6: Previous batch positive control (10^5^ per μl). Tube 7: MERS-CoV DNA (negative control), 10^4^ per μl. Tube 8: No template control (NTC).

## Discussion

In this study, we confirm that one of the fastest, simplest, cheapest and most scalable protocols available for detecting SARS-CoV-2 nucleic acid is so far reliable for strong positives only, but highly specific compared to “gold standard” qRT-PCR detection. Other studies have shown similar reduced sensitivity but high specificity ^**17-21**^. One early study found that RT-LAMP was more sensitive than RT-PCR at detecting virus in an asymptomatic carrier monitored for several days ^**22**^. RT-LAMP has been tested extensively at large scale in the UK and has recently been shown to have less-than-expected sensitivity (<50%) when rolled out to large populations as part of Operation Moonshot ^**23**^. However, similarly to the current study, sensitivity was greatest in strong positives, indicating that only those with high viral loads may reliably be detected with this diagnostic platform, especially when performed by non-experts in field settings.

The pace of Canadian approvals for SARS-CoV-2 point-of-care diagnostic tests has been much faster than for other infections over the years (such as HIV/STBBI). Initial conditional approvals were revoked for the isothermal amplification-based Spartan Cube and its apparently inadequate silicon sampling devices. Recent approvals have included a rapid isothermal platform (Abbott’s IDNow, based on nicking enzyme amplification reactions or NEAR) and a rapid antigen-detecting cassette (Abbott’s PanBio Ag Rapid Test Device), adding to the successful Xpert® Xpress SARS-CoV-2 (Cepheid®). No rapid, point-of-care antibody tests have yet been approved for use in Canada.

The FDA has meanwhile provided Emergency Use Authorization for dozens of diagnostics of all descriptions, from the Sherlock Biosciences LAMP/CRISPR platform to the recent LAMP-based home test by Lucira ^**24**^. All of these tests are imperfect and do not work as well in practice as manufacturer’s claim. Despite these limitations, all tests applied on a wider scale, with proper messaging about the limited significance of negative results, could be highly useful if strong positives are identified and acted upon more quickly ^**25**^. For example, several jurisdictions are exploring use of PanBio Ag Rapid Test Device to identify positives early, assuming that negative results will need additional testing in order to be confirmed ^**26-28**^.

Molecular diagnostic techniques that use raw or minimally processed sample are fastest, but less sensitive, since the background chemistry of a raw sample may introduce uncontrolled variability ^**18,29**^. Potential inhibitors or other contaminants may reduce the reliability of both positive and negative results. This concern is somewhat lessened in the context of the current report, which we have confirmed is highly specific and can amplify target molecules even in highly diluted samples. However, the current assay’s limit of detection (currently 1,000 copies/μl, similar to other studies ^**18**^) does decrease the likelihood of detecting a person with low viral load. In contrast, Sherlock Biosciences claims on its website (www.sherlock.bio) that its LAMP-based assay can detect 7 copies per μl VTM or up to 150 times less than this report.

Is the risk of missing someone with a low viral load greater than making a person with a high viral load wait days to find out they are positive? In all cases, expectations about what a negative test result *actually means* must be properly managed. In this study, RT-LAMP with raw, heat-treated sample was ∼75% sensitive compared to nasopharyngeal RT-PCR (“the gold standard”), meaning that a false negative result can be expected 1 out of 4 times the test is performed. Therefore, careful explanation that a single negative test does *not* mean a person is free of SARS-CoV-2 must be included as part of pre-test and post-test counselling.

However, RT-LAMP using raw sample is much cheaper and faster than RT-PCR using purified RNA (Table 2). Multiple RT-LAMP tests can be conducted in less time and at less cost than a single “gold standard” RT-PCR test. Repeat screening over several days may also help identify the earliest timepoint at which a person is infectious, increasing confidence that a negative test result is not just due to random error or low test sensitivity ^**30**^, and increasing the impact of contact tracing.

**Table 2:** Base cost (CAD$) of commercial kit and procedure time to conduct extract-free LAMP assay.

|  | per reaction | per result<sup>1</sup> | per 24 | per 96 |
| --- | --- | --- | --- | --- |
| **WarmStart Colorimetric Kits<sup>2</sup>** | ~\$2-3 | ~\$10-15 | ~\$150-\$230 | ~\$600-\$900 |
| 2X Master Mix | \$1.76-2.82 | \$8.45-13.53 | \$135-216 | \$540-860 |
| LAMP Primer Mixes | \$0.02-0.06 | \$0.10-0.30 | \$2.40-7.20 | \$10-\$30 |
| **Procedure time<sup>3</sup>** | - | ~1-1.5 hours | 2-3 hours | 3-5 hours |
| Master mix setup (96) | - | 4 min. | 15 min. | 30 min. |
| Sample processing/dilution | - | 15 min. | 60 min. | 90 min. |
| Transfer to Master Mix | - | 5 min. | 15 min. | 30 min. |
| Isothermal amplification | - | 35 min. | 35 min. | 35 min. |
| Evaluate and record result | - | 6 min. | 15 min. | 45 min. |
| Repeat failed reactions | - | 45 min. | 65 min. | 110 min. |

<sup>1</sup> Assuming an average of four reactions per screen (multiple dilutions and/or primer sets), plus 20% for cost of positive and negative controls (4 per 48 reactions) and re-tests (6 per 48 reactions). <sup>2</sup> Cost variance due to different sizes of kits and formulations available and whether 20 or 25 µl test volume is used. <sup>3</sup> Does not include sample collection or transport time.

Ideally, new testing modalities should not compete with gold standard tests for equipment, consumables or personnel, all of which are presently stretched very thin. Provincial laboratories and diagnostic services in hospitals are diverting high throughput platforms in attempt to keep up with surges in testing for SARS-CoV-2 at the expense of testing for other pathogens. Developing multiple testing platforms that do not require the same instruments, reagents and laboratory infrastructure are of paramount importance in order to increase testing capacity ^**25**^. RT-LAMP uses Bst instead of Taq for amplification, does not compete for tips and reagents for RNA extraction (if raw sample is used), and is simple enough for a trained non-specialist to execute with oversight and support, freeing nurses and other front-line staff for less technical duties. Increased validation and application of rapid tests such as RT-LAMP is only worthwhile if it frees up central laboratories to get back to providing diagnostics services, of all kinds, to those most in need.

In conclusion, scalable rapid tests, such as the one evaluated in this study, may efficiently detect individuals with high viral loads at the point of care. Our findings suggest RT-LAMP could be useful as a screening mechanism for prioritized samples within the existing test chain, reliably identifying those with highest virus concentrations hours ahead of the standard RT-PCR workflow, and able to be scaled to any required number of tests per day. Further work must focus on improving sensitivity, incorporating saliva or other self-collected samples, and triangulating evidence from different testing modalities ^**31**^, to better ascertain an individual’s infectious period.

## Data Availability

All data is included in the manuscript.

## Acknowledgements

Thanks to Dr. Kerry Dust, Dr. Paul van Caeseele, Dr. Jared Bullard, Dr. Derek Stein, Adam Hedley and all the staff and scientists at Cadham Provincial Laboratory who originally processed the samples for SARS-CoV-2 testing. Thanks to Dr. Keith Fowke, Dr. Grant McClarty, Dr. Pam Orr, Sheila Ang and Steve Wayne from the Department of Medical Microbiology and Infectious Diseases, Rady Faculty of Health Sciences, University of Manitoba, for space, advice, encouragement and practical support.

